# Peak Agricultural Pesticide Spray Periods are related to Real-World Cognition Using an Ecological Momentary Assessment Burst Design over Two Years

**DOI:** 10.64898/2026.11.02.26367130

**Authors:** Briana N.C. Chronister, Emma Parrish, Rajendra Parajuli, Dolores Lopez-Paredes, Harvey Checkoway, Jose Suarez-Torres, Avery Quynh, Danilo Martinez, Asa Bradman, Jose R. Suarez-Lopez, Raeanne C. Moore

## Abstract

**Objective:** Emerging evidence suggests that exposure to organophosphate, neonicotinoid, and other pesticides may transiently impair cognition; however, traditional "snapshot" laboratory-based cognitive assessments fail to capture fluctuations in cognitive performance. This study used ecological momentary cognitive testing (EMCT) to evaluate whether real-world cognitive performance differs between peak and low floricultural pesticide spray seasons, thereby evaluating the potential cyclical effects of agricultural production and related pesticide exposures.

**Material and Methods:** From 2022 to 2024, 15,063 EMCT assessments were completed by 494 participants (ages 17–25; 50% female) enrolled in the ESPINA cohort study in Ecuador.

Participants completed nine 10-day burst assessments every 2–3 months during high and low pesticide spray seasons. EMCTs (NeuroUx) assessed processing speed, executive function, and visual working memory. Mixed-effects models examined associations of peak versus off-peak pesticide spray seasons, floricultural work or occupation (vs not) with EMCT performance, adjusting for socio-economic and other variables, and including participant- and burst-level random intercepts.

**Results:** Peak pesticide spray season, compared to off-peak, was associated with higher T-scores on processing speed tests, including Matching Pair (β=0.62) and Odd One Out (β=0.88), but showed no significant association with other tests or the overall cognitive composite. Conversely, direct floricultural employment was significantly associated with lower cognitive performance overall (β=−0.59), with the largest differences observed in Memory Matrix (β=−0.78) and Matching Pair (β=−0.70).

**Conclusion:** Peak pesticide spray seasons were associated with modestly higher processing speed scores, consistent with prior findings in this population suggestive of enhanced cognitive performance associated with neonicotinoid and sulfoximine exposures. In contrast, floricultural employment was associated with significantly lower memory and overall cognitive performance. These results suggest that occupational and seasonal pesticide exposure may have distinct associations with cognitive functioning, highlighting the importance of repeated cognitive testing for characterizing potentially transient effects.

## Introduction

Emerging evidence suggests that cognitive health is shaped not only by social and behavioral determinants but can also be impacted by environmental exposures^1^. Among these, pesticide exposure has become a growing concern in agricultural communities, where compared to individuals living in urban regions, agricultural populations may experience repeated, low-level contact with complex mixtures of pesticides through occupational tasks, para-occupational pathways, contaminated household environments, and pesticide drift from nearby crop production. Although pesticides are designed to target biological systems in pests, many widely used compounds, including organophosphates, pyrethroids, neonicotinoids, and herbicides, have also been implicated in pathways relevant to human neurotoxicity, including cholinergic dysregulation^2^, oxidative stress^3^, mitochondrial dysfunction^4^, inflammation^5,6^, and endocrine disruption^7–9^.

Mounting evidence has linked pesticide exposure to adverse neurobehavioral outcomes, including poorer neurocognitive performance. For example, farmers in Southern France with direct or indirect pesticide exposure were at greater risk of poor performance across multiple neurobehavioral measures, including the Mini-Mental State Examination, Benton Visual Retention Test, Trail Making Test Part A, and Stroop Test^10^. Although pesticide classifications act through different biological mechanisms, exposure across several major pesticide types have been associated with worse neurobehavioral outcomes across various populations^11–14^. In our prior work in Ecuador, organophosphate exposure has been linked to poorer neurobehavioral performance in children, with reduced acetylcholinesterase activity associated with lower NEPSY-II performance in attention and inhibitory control, visuospatial processing, and sensorimotor domains^15^. Among adolescents, exposure to herbicides 2,4-D and glyphosate was associated with poorer performance in attention and inhibitory control, memory and learning, and language^16^. In contrast, we also previously observed some evidence that urinary neonicotinoid metabolites were associated with greater performance in social perception and memory in adolescence and again in young-adulthood, consistent with the short-term enhancing properties of nicotinic acetylcholine receptor agonists^17^. Another pediatric population recently reported greater behaviors of hyperactivity among children with greater urinary neonicotinoid metabolites^18^. Pyrethroid insecticides have also been associated with neurocognitive alterations, particularly in attention, processing speed, and motor coordination^19^. Collectively, these findings indicate that exposure to commonly used agricultural pesticides may disrupt neurobehavioral functioning across multiple stages of development.

Floricultural and agricultural occupations represent one of the most sustained routes of pesticide exposure, yet their independent associations with cognitive performance remain relatively understudied compared with the broader biomarker literature. A systematic review, focused on 106 articles on brain health in agricultural workers, further identified pesticides as the most studied occupational risk factor, with agricultural work associated with alterations in dopaminergic and cholinergic neurotransmitter systems and disrupted functional brain connectivity^20^. Workers engaged in floriculture encounter complex pesticide mixtures repeatedly across planting, cultivating, and harvesting cycles^21^. Evidence from farmworker populations further supports cognitive effects associated with occupational pesticide exposure. Mexican avocado farmworkers with higher urinary concentrations of organophosphate and pyrethroid metabolite performed worse on measures of processing speed and executive function^22^. Meta-analytic evidence similarly indicates that even low-level occupational organophosphate exposure is associated with impairment across multiple cognitive domains^23^.

Pesticide exposure does not end at the worksite. Family members of floricultural workers may experience secondary exposure through contaminated work clothing, household dust, and pesticide drift^24–26^. Residential proximity to greenhouse floriculture has been associated with elevated urinary pesticide metabolite concentrations in adolescents^27^, and children living closer to agricultural land showed worse neurobehavioral performance in attention, language, and memory^28^. Together, these findings establish that floricultural occupation and household cohabitation with agricultural workers are distinct but overlapping exposure pathways, each carrying neurocognitive implications for workers and those who share their living environments.

In agricultural communities, pesticide exposure is also not static. Exposure intensity may vary across the year in relation to crop cycles, pest pressure, and commercial production demands, creating recurring periods during which residents may experience heightened environmental exposure^29–31^. These temporally patterned exposure windows may be especially relevant for identifying short-term changes in cognition, yet they remain poorly characterized in epidemiologic research. Preliminary evidence from the Study of Secondary Exposures to Pesticides Across Childhood, Adolescence, and Adulthood (ESPINA) suggests that children’s neurobehavioral performance recovers following peak pesticide use periods, potentially returning to baseline as exposure drops^15,32^. Traditional study designs often lack the temporal resolution necessary to examine whether cognition differs during periods of peak agricultural pesticide spraying compared with lower-spray periods.

Most studies linking pesticide exposure to neurobehavioral outcomes have relied on cross-sectional designs, single assessments, or clinic-based testing that may not capture real-world cognitive variability. This limitation is particularly relevant during adolescence and young adulthood, when executive function, working memory, processing speed, and attention continue to mature, shaping learning, emotional regulation, and long-term functioning^33–37^. Ecological momentary cognitive testing (EMCT) addresses this gap through repeated, near real-time assessments in natural environments, improving ecological validity and capturing within-person variability across changing environmental conditions^38^. Repeated burst designs are particularly suited to examining seasonal pesticide exposures and associated cognitive changes.

To our knowledge, no prior study has used longitudinal EMCT to examine cognitive performance across peak and low pesticide spray periods in agricultural communities or evaluated the independent associations of floricultural employment and household cohabitation with agricultural workers over multiple years. Addressing these gaps could identify modifiable environmental determinants of cognitive health in agricultural populations.

In this study, we used an EMCT burst design over two years within the ESPINA cohort to evaluate whether peak agricultural pesticide spray periods were associated with real-world cognitive performance among adolescents and young adults living in an agricultural community in Ecuador. We hypothesized that cognitive functioning would be poorer during peak versus low pesticide spray periods and among participants with greater occupational or household pesticide exposure compared with unexposed counterparts.

## Methods

### Study Design

ESPINA-EMCT is an ancillary study that was launched in tandem with the 14th year follow-up year (FUY) the ESPINA study in July 2022 (FUY-14a), in Pedro Moncayo Canton, Ecuador. Enrollment details into the original ESPINA cohort have been previously published^17,26,39^. In summary, ESPINA is a longitudinal cohort study that has conducted 13 follow-up examinations over 16 years since its inception in 2008 when participants were 4-7 years old. Since 2008, ESPINA has examined 751 participants from childhood through young adulthood to assess the health impacts of pesticide exposure across the life course^26,40,41^.

Participants have been recruited from the Survey of Access and Demand of Health Services (SADHS), the System of Local and Community Information (SILC), community announcements, and word-of-mouth.

ESPINA-EMCT included one completed in-person assessment and nine 10-day bursts once every 2-4 months from July 2022 (FUY-14a) to September 2024 (FUY-16d). For the FUY-14a assessment, 386 participants had completed the ESPINA FUY-8b assessment, and 119 participants were newly recruited using the SILC, community announcements, and word-of-mouth like prior assessments.

### Setting

Pedro Moncayo, Ecuador has a population of approximately 33,000 and is known for its extensive floricultural production, primarily consisting of roses and some other ornamental flowers^42,43^. In 2022, floricultural greenhouses comprised ∼5% of Pedro Moncayo’s total surface area. Favorable environmental conditions, including minimal seasonal variation in temperature, precipitation levels, day length, and fertile soil, supports continuous crop cultivation throughout the year^42–44^. Roses are perennial flowers^45^ with peak harvest periods in Ecuador corresponding to major international holidays, including New Year’s in January, Valentine’s day in February, Mother’s Day, which is celebrated in May in ∼63% of the world^15^, and Christmas in December. To satisfy increased export demands during these periods, pesticide application intensifies, resulting in harvest seasons that overlap with peak pesticide spraying seasons.

### Ecological Momentary Cognitive Testing (EMCT)

Participants completed a brief EMA survey alongside a suite of six mobile cognitive tests in Spanish using NeuroUX, a digital health platform that has been used across diverse populations^46,47^. The mobile survey and all test stimuli (including instructions) for the mobile tests were translated from English into Spanish by members of the research team, which included people native to Pedro Moncayo County to ensure that the Spanish terminology used was appropriate for the region. The first EMCT administration was conducted in-person during the FUY-14a examination at study sites in Pedro Moncayo, using study-provided smartphones. Subsequent administrations were conducted remotely using participants’ personal smartphones through a NeuroUX app, with assessments completed daily during administrations 2–4 and every other day during administrations 5–10. Participants completed these assessments in their daily environments and were instructed to select times and settings with minimal distractions. Each EMCT session, which included the mobile tests and a brief survey, averaged 12.5 minutes. Validation of the mobile cognitive tests in this study population have previously been published^38^. The following tests were administered and are briefly described below (additional details have been published previously^38,46,48^):

***Quick Tap 1*** (Domains Assessed: Processing Speed and Reaction Time). Participants respond as quickly as possible to a visual target presented on the screen. Performance is characterized by response accuracy and reaction time.

***Quick Tap 2*** (Domain Assessed: Executive Function). Participants respond to target stimuli while inhibiting responses to non-target stimuli. Performance is characterized by response accuracy and reaction time.

***CopyKat*** (Domain Assessed: Visual Working Memory). Participants reproduce increasingly complex visual sequences presented on the screen. Performance is characterized by accuracy.

***Memory Matrix*** (Domain Assessed: Visual Working Memory). Participants recall and identify the locations of visual stimuli presented within a matrix, with increasing difficulty across correct trials. Performance is characterized by recall accuracy.

***Odd One Out*** (Domains Assessed: Processing Speed and Reaction Time). Participants identify and select the unique stimulus among a set of similar visual stimuli. Performance is characterized by accuracy and reaction time.

***Matching Pair*** (Domain Assessed: Processing Speed). Participants identify matching visual stimuli presented within a matrix, which increases in size across correct trials.

Performance is characterized by the number of correctly completed items within the allotted time.

### NeuroUX Total Score Creation

Total scores for Memory Matrix, Matching Pair, and Odd One Out were equal to their raw scores. Quick Tap 1, Quick Tap 2, and Odd One Out Total Scores were revised using a two-vector scoring method based on the accuracy and reaction time, similar to the Toolbox FICA Test and Dimensional Change Card Sort Measures, as previously published ^38^. In brief, Quick Tap 1, Quick Tap 2, and Odd One Out total scores were calculated using weighted formulas combining reaction time and accuracy components, with weights of 90% time and 10% accuracy for Quick Tap 1 and 60% time and 40% accuracy for both Quick Tap 2 and Odd One out. Accuracy and reaction-time components were scaled separately and combined into total scores ranging from 0 to 10, with higher scores indicating better performance. Accuracy weights differed by test to account for ceiling effects and score distributions, while reaction times were log-transformed, bounded by test-specific minimum and maximum values, and reverse-scored so that faster responses received higher scores.

### NeuroUX T-Score Creation and Composite Score

All total scores were converted to T-scores, adjusting for race, age, gender, and education status. The composite cognitive score averaged the T-scores across all tests, excluding Odd One Out due to day-to-day variability in task difficulty and CopyKat due to participant-reported reductions in effort at higher performance levels, attributed to the length of the test.

### Participant Demographics

Surveys of participant characteristics were conducted at FUY14a, 16a and 16c. To maximize observations, a rule-based single imputation strategy that leveraged repeated measures from ESPINA-collected survey information was used. To determine average parental education, we averaged the participant-reported maternal and paternal years of education from FUY-14a. If unavailable, we imputed the value based on reported parental education from the FUY-8b, then we used participant-reported parental education from the NIH Toolbox captured in FUY-14a. For parents classified in the lowest NIH education category (“no schooling”), we assigned 0 years of education. When all sources were missing, we imputed the sample mean (9.22 years). Missing data for monthly household income, occupation, and cohabitation status was imputed sequentially using data reported from the proceeding assessment (e.g., FUY-14b imputed using FUY-14a values). If covariate data was missing at all time points for categorical variables, those variables were recategorized as “Missing/Prefer not to answer.”

### Statistical Analysis

Descriptive statistics were calculated, and differences between high and low pesticide spray seasons were assessed using independent t-tests for continuous variables, or chi-squared analyses for categorical variables. Generalized linear mixed effects models (GLMM), with a participant level and nested burst random intercept, were implemented to assess the association between high pesticide spray season (versus low pesticide spray season) with mood outcomes, including domain-level (e.g., positive affect) and individual symptoms (e.g. happiness) scores. Fixed effects included day of assessment (1–10) and a set of a priori–selected confounders: age, sex, monthly household income, participant education, average parental education, employment as a floricultural worker, and cohabitation with a floricultural worker.

Burst 1 EMCTs varied from Bursts 2 to 10, as Burst 1 was completed the same day as the participants in-person visit, while the EMCTs for the remaining bursts occurred in real-world settings. Thus, we also included a clinic covariate to control for differences in completion of the EMCTS in effect modification by sex was assessed by introducing an interaction term between pesticide spray season and sex. If the p<0.05, then the GLMM was stratified by sex. Lastly, to assess within-person effects, we used linear mixed models centered on each participant’s mean value for each of the EMA variables, based on the mean values for participant’s own mean values. All analyses were performed using R, version 2025.09.2.

## Results

A total of 489 participants contributed 13,681 observations (on average 28 assessments per participant), including 4,745 observations during low pesticide spray seasons and 8,936 during high pesticide spray seasons (Table 1). Participants had a mean age of 20.3 years (SD = 1.8), and 50.5% of unique participants were female. A comparison of unique individuals to total observations reveals that certain demographics were significantly more likely to consistently complete EMCT assessments. Most notably, females demonstrated the highest compliance, making up just 50.5% of the enrolled cohort but generating 61.7% of the total data points.

**Table 1.** Participant Demographics of unique participants, and total observations for the ESPINA-EMCT cohort.

|  | **Unique Individuals** | **Total Observations** | **Low Pesticide Spray Season** | **High Pesticide Spray Season** | **p-diff** |
|---|---|---|---|---|---|
| **Observations** | 440 | 13681 | 4745 | 8936 |  |
| **Age (years)** | 20.97 (1.86) | 21.37 (1.92) | 21.68 (1.94) | 21.21 (1.89) | <0.001 |
| **Female (%)** | 52.6% | 61.7% | 60.2% | 62.5% | 0.01 |
| **Race** |  |  |  |  | 0.42 |
| Indigenous | 16.1% | 13.8% | 14.1% | 13.6% |  |
| Mestizo, White or Other | 83.9% | 86.2% | 85.9% | 86.4% |  |
| **Monthly Household Income (%)** |  |  |  |  | 0.38 |
| Less than \$500 | 52.3% | 52.7% | 52.2% | 53.0% |  |
| \$500-\$999 | 23.2% | 23.9% | 24.5% | 23.5% |  |
| \$1000+ | 5.9% | 4.8% | 5.1% | 4.7% |  |
| Prefer not to Answer | 18.6% | 18.6% | 18.3% | 18.8% |  |
| **Average Parental Education (years)** | 9.11 (3.64) | 9.10 (3.60) | 9.18 (3.64) | 9.06 (3.58) | 0.06 |
| **Participant Education** |  |  |  |  |  |
| Elementary and Middle School | 2.0% | 1.0% | 1.0% | 1.0% | 0.91 |
| Some High School | 12.0% | 11.5% | 11.4% | 11.6% |  |
| High School Graduate | 44.8% | 42.2% | 42.6% | 42.0% |  |
| Some College to College Grad | 41.1% | 45.3% | 45.0% | 45.5% |  |
| **Works in Floriculture (%)** | 17.0% | 14.1% | 14.9% | 13.7% | 0.07 |
| **Lives with a Floricultural Worker (%)** | 58.6% | 59.4% | 59.0% | 59.5% | 0.57 |
| **AChE** | 4.11 (0.52) | 4.04 (0.58) | 4.11 (0.59) | 3.96 (0.56) | <0.001 |
| **Cumulative Tests Taken** | - | 20.4 (14.9) | 27.1 (16.2) | 16.9 (12.8) | <0.001 |
| **Time of the Assessment** |  |  |  |  | <0.001 |
| Before 8am | - | 8.8% | 10.3% | 7.9% |  |
| 8am to 8 pm | - | 79.7% | 76.3% | 81.5% |  |
| After 8 pm | - | 11.5% | 13.3% | 10.5% |  |
| **Weekend Assessment** | - | 19.2% | 20.4% | 18.5% | 0.01 |
| **Alone During Assessment** | - | 41.5% | 39.1% | 42.8% | <0.001 |
| **At Home During Assessment** | - | 75.0% | 70.7% | 77.3% | <0.001 |

p-diff was calculated using chi-square for categorical data, and ANOVA for continuous variables.

Participants with at least some college education and those who were slightly older also completed a disproportionately higher number of assessments. Conversely, direct employment in the floriculture industry appeared to hinder ongoing participation; floriculture workers comprised 18.8% of the unique individuals but only 14.1% of the total observations. Indigenous participants similarly contributed fewer assessments per capita than their non-indigenous peers. AChE activity was significantly lower during high-spray than low-spray seasons (Table 1), providing an objective biological support for the predefined seasonal classification.

The number of EMCT observations differed across pesticide spray seasons, with more assessments completed during high-spray (n = 8,936) than low-spray (n = 4,745) seasons.

Assessment context also varied by season. During high-spray seasons, participants were more likely to complete assessments at home (77.3%), alone (42.8%), on weekdays, and during daytime hours (8 AM–8 PM; all *p* < .001).

Across all observations, the overall mean NeuroUX Composite Score was 50.62 (SD = 5.36). Performance on the individual subtests was highly consistent, with average T-scores closely anchored to 50. This central tendency was expected, as the T-scores were created and standardized based directly on the ESPINA-EMCT study’s own observations. Specifically, participants scored an average of 50.04 on both the Memory Matrix (SD = 10.58) and CopyKat (SD = 8.88) tasks, 50.01 on Quick Tap 2 (SD = 10.72), 49.98 on both Matching Pair (SD = 10.57) and Quick Tap 1 (SD = 9.74), and 49.96 on Odd One Out (SD = 11.38).

### Cognitive Performance Across Bursts

Cognitive performance demonstrated significant longitudinal trends across bursts, although the direction and magnitude varied by test (Figure 1). The NeuroUX composite scores increased across successive bursts (β = 0.030; 95% CI: 0.029, 0.030). Small positive longitudinal trends were also observed for Memory Matrix (β = 0.008; 95% CI: 0.008, 0.009), CopyKat (β = 0.007; 95% CI: 0.007, 0.008), and Matching Pair (β = 0.003; 95% CI: 0.002, 0.003). In contrast, performance declined across successive bursts, with the largest decrease observed for Odd One Out (β = −0.022; 95% CI: −0.022, −0.021), followed by Quick Tap 2 (β = −0.017; 95% CI: −0.017, −0.016) and Quick Tap 1 (β = −0.009; 95% CI: −0.010, −0.009).

**Figure 1.**
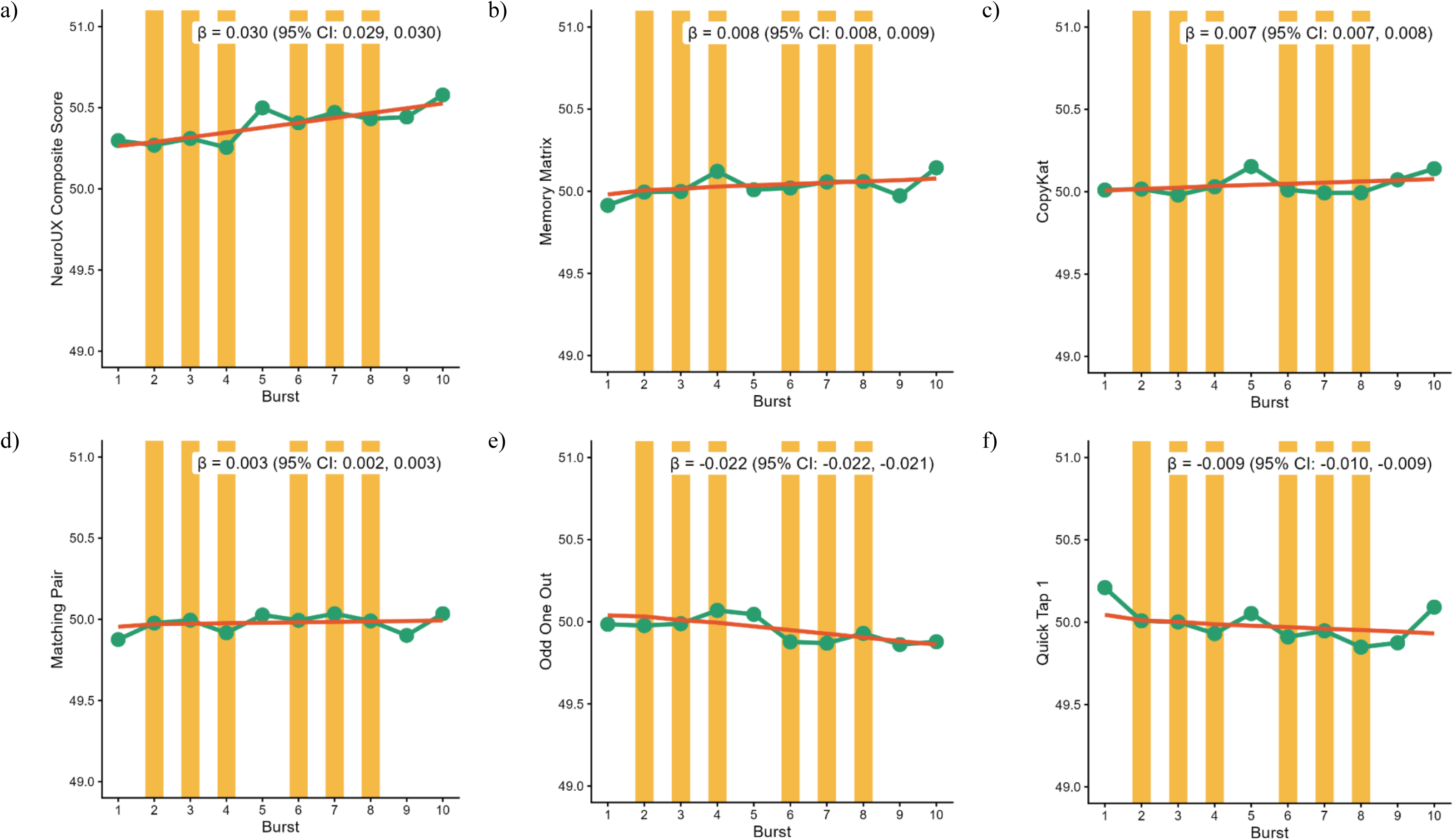
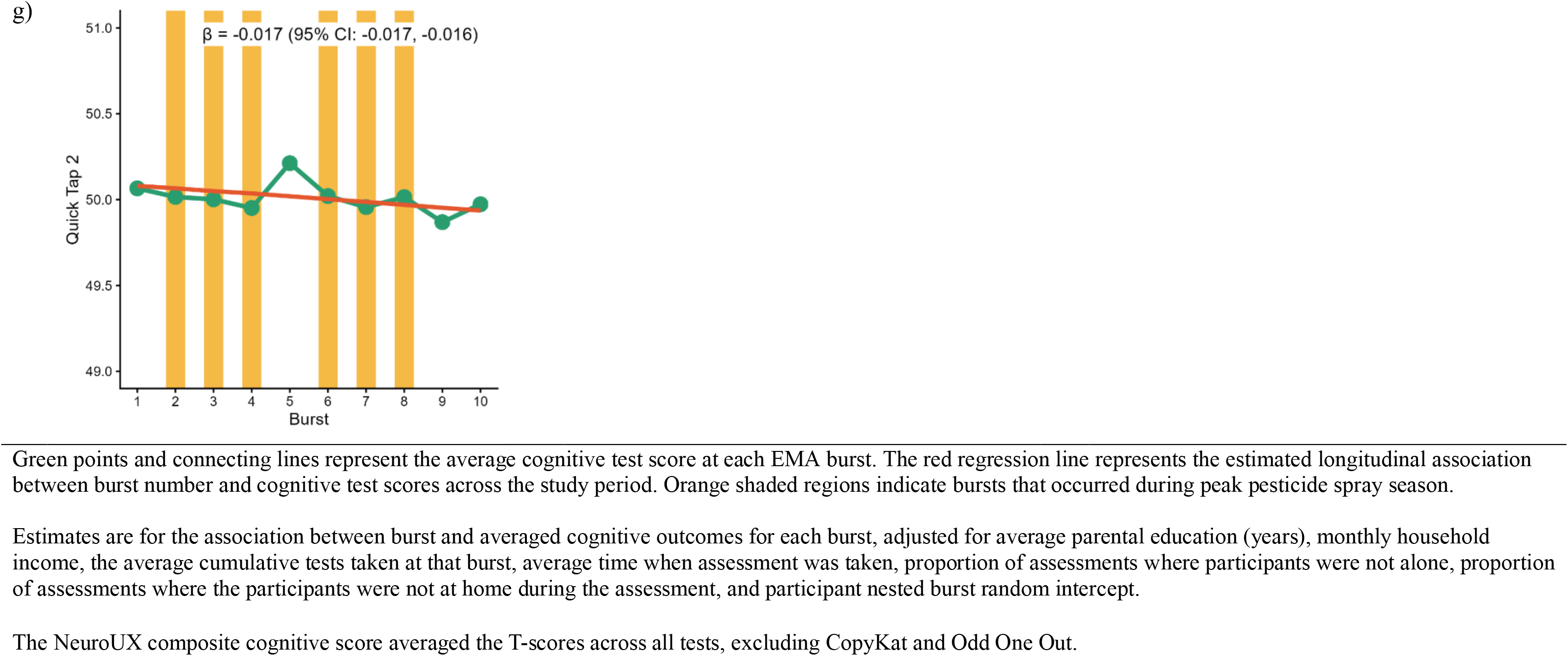
Mean NeuroUX scores by bursts and longitudinal association between burst number and test performance for ESPINA-EMCT (n=489, nobs=13,681) from July 2022 to September 2024.

### Seasonality with Cognitive Performance

Peak pesticide spray season, versus off-peak, was associated with higher T-scores on processing speed tests including Matching Pair (β =0.62 [0.18, 1.05]) and Odd One Out (β =0.88 [0.40, 1.35]) (Figure 2), but no significant differences were observed for Memory Matrix (β =0.25 [−0.20, 0.70]), Quick Tap 1 (β =−0.05 [−0.47, 0.38]), Quick Tap 2 (β =0.19 [−0.26, 0.65]), or CopyKat (β =−0.23 [−0.67, 0.22]). Similarly, there was no significant association between peak pesticide spray season and the overall NeuroUX cognitive composite (β =0.16 [−0.12, 0.44]). We did not observe interaction effects between peak spray season with gender, floricultural occupational status, nor cohabitation with a floricultural worker, thus the GLMM models were not stratified.

**Figure 2.**
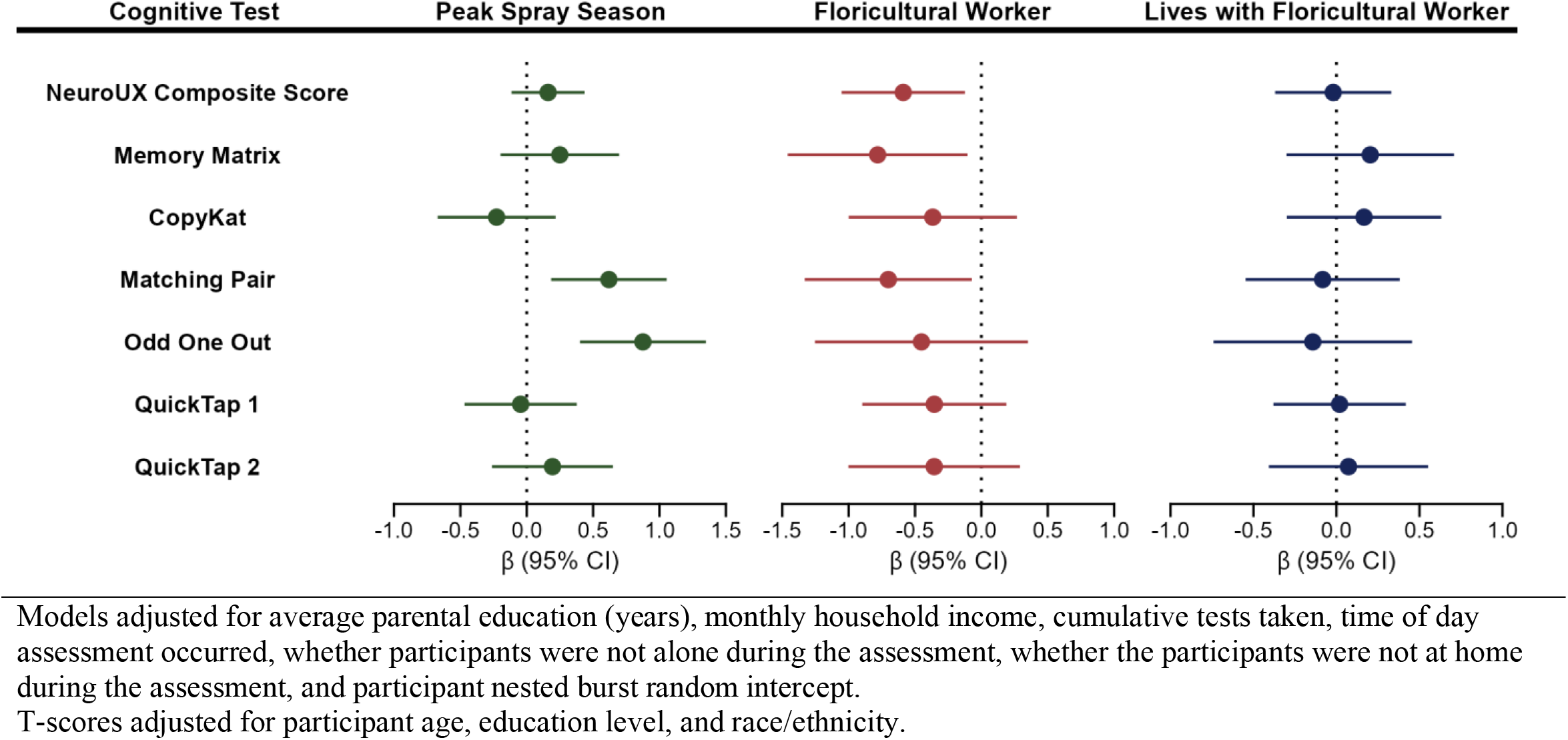
Differences in ecological momentary cognitive test T-scores during peak versus off-peak pesticide spray seasons, and among participants with personal or household occupational exposure to floriculture versus unexposed participants (ESPINA-EMCT study, July 2022–September 2024; n=489, nobs=13,681).

### Floricultural Work and Cohabitation with Floricultural Workers in Relation to Cognitive Performance

Floricultural workers generally had lower cognitive performance than non-floricultural workers in longitudinal models after adjustment for socioeconomic and other variables (Figure 2). The largest negative associations were observed for the NeuroUX Composite Score (T-score difference [β]=−0.59 [−1.05, −0.12]), Memory Matrix (β=-0.78 95%CI: [-1.46, -0.10]), and Matching Pair (β=-0.70 95%CI: [-1.32, -0.07]. Associations for QuickTap 1, QuickTap 2, Odd One Out, and CopyKat were also negative, although not statistically significant. Cohabitation with a floricultural worker was not associated with changes in cognitive performance (Figure 2). We did not observe effect modification by gender for floricultural work or cohabitation with floricultural workers in relation to cognitive performance.

## Discussion

In this longitudinal investigation of the ESPINA-EMCT cohort, we assessed the impacts of seasonal and occupational agricultural exposures on the cognitive performance of young adults using high-frequency ecological momentary cognitive testing over two years. Our primary findings reveal a stark divergence in cognitive outcomes depending on the nature of the exposure. Direct employment in the floriculture industry was consistently associated with significantly lower overall cognitive performance, particularly in working memory and processing speed. However, these negative impacts appear localized to direct occupational hazards, as indirect household exposure (measured via cohabitation with a floricultural worker) yielded no significant cognitive differences. Furthermore, we observed an unexpected seasonal dynamic: assessments completed during peak floricultural pesticide spray seasons were associated with higher processing speed scores rather than lower performance. These differences reflex moderate variations in cognitive performance within a cognitively normal young adult sample, rather than clinically meaningful impairment. Together, these findings suggest that while chronic occupational exposure presents cognitive risks, specific seasonal pesticide exposures may transiently stimulate certain cognitive domains.

The positive associations observed between peak pesticide spray season and performance on processing-speed tasks (Matching Pair and Odd One Out) must be interpreted cautiously. This seasonal enhancement aligns with prior observations in this cohort, where adolescent and young adult exposures to specific agricultural chemicals (notably neonicotinoids and sulfoximines) were linked to greater cognitive performance^49,50^. A plausible mechanism involves the structural and functional similarities between neonicotinoid insecticides and nicotine; both excite nicotinic acetylcholine receptors^51^, a pathway known to mediate the short-term enhancing effects of nicotine on memory and attention^52–55^. While we have previously found exposures to organophosphates and, to some extent, pyrethroids were associated with lower neurocognitive performance^49,50,56^, agricultural practices have shifted over time. As organophosphate use has declined and neonicotinoids have increasingly replaced them, the "peak pesticide spray seasons" captured between 2022 and 2024 may more strongly reflect the stimulatory effects of neonicotinoids than the detrimental impacts of older chemical classes.

Nevertheless, these results should not be interpreted as evidence of a net beneficial effect of peak pesticide application periods. Peak spray season serves only as a broad, indirect proxy, and actual individual pesticide burdens vary substantially by crop type, producer, and application timing. Furthermore, spray periods coincide with a host of other seasonal changes that are known to influence cognition, including shifts in work and school schedules, sleep patterns, social activity, and environmental conditions. For instance, peak agricultural periods often demand increased overtime work among floricultural workers; a factor previously associated with poorer attention and executive function among industrial workers^57^. Therefore, the observed seasonal associations likely reflect a complex interplay of specific, evolving chemical exposures and concurrent contextual factors that fluctuate across the agricultural cycle. The lower AChE activity observed during high-spray seasons provides some biological support for the distinction between the predefined spray seasons. However, because AChE primarily reflects exposure to cholinesterase-inhibiting pesticides, it does not quantify exposure to the broader mixture of pesticides used in floriculture. Future work incorporating biomarkers of multiple pesticide classes is needed to better characterize associations between seasonal pesticide exposure and cognitive performance.

The repeated nature of the NeuroUX assessments is also important when interpreting these seasonal findings. Cognitive performance changed significantly across successive assessment bursts, with the direction of these longitudinal trends varying by task. Performance increased over time for the overall NeuroUX Composite Score, Memory Matrix, CopyKat, and Matching Pair, whereas performance declined for Odd One Out, Quick Tap 1, and Quick Tap 2. These heterogeneous trajectories may reflect task-specific combinations of practice effects, familiarity with the mobile testing platform, changes in participant engagement, learning, or fatigue. Repeated cognitive testing can result in improved performance as participants become increasingly familiar with task instructions and response requirements, although the magnitude of such effects can differ substantially across cognitive domains. Conversely, declining performance on some tasks may indicate reduced engagement or motivation over repeated administrations. Because high and low pesticide spray seasons occurred at different points throughout follow-up, accounting for repeated testing and temporal trends (accounted for in this manuscript) is particularly important when estimating seasonal associations.

In contrast to the partial transient seasonal enhancements, floricultural employment was associated with modestly lower cognitive performance across the battery. Floricultural workers had significantly lower Memory Matrix and Matching Pair scores and a lower overall NeuroUX Composite Score than participants who were not employed in floriculture. However, scores in both groups remained within the average range of performance, indicating relative differences in performance rather than clinically meaningful cognitive change. Associations for all remaining cognitive tests were also in the direction of poorer performance among floricultural workers, although their confidence intervals included zero. The consistency in the direction of these associations strengthens the possibility that occupational circumstances contribute to cognitive differences even when individual task-specific associations do not reach statistical significance.

These occupational findings are broadly consistent with concerns regarding the potential neurobehavioral consequences of agricultural pesticide exposure. Floricultural workers may experience more direct or frequent pesticide exposure through contact with treated plants or surfaces and proximity to pesticide applications than individuals whose exposure arises primarily from living in an agricultural community. However, floricultural employment should not be interpreted solely as a measure of pesticide exposure. Employment in this industry may capture pesticide exposure alongside numerous other occupational characteristics, including physical workload, working hours, heat exposure, socioeconomic conditions, stress, and other workplace exposures. The observed associations therefore cannot establish that pesticide exposure itself accounts for poorer cognitive performance among floricultural workers. Future studies incorporating biomarkers of pesticide exposure, detailed occupational histories, and information on specific work activities will be important for determining which aspects of floricultural employment contribute to these cognitive differences.

The absence of associations between cohabitation with a floricultural worker and cognitive performance provides important context for the occupational findings. The absence of cognitive associations among participants cohabitating with floricultural workers provides important context for our occupational findings. Although household members may experience indirect, para-occupational pesticide exposure via contaminated clothing or equipment, this contact is substantially lower than direct occupational exposure. The contrast between cognitive differences in floricultural workers and null findings in their housemates suggests an exposure gradient, where direct occupational hazards confer greater neurobehavioral risks. However, without direct exposure measurements, these differences cannot be definitively attributed to varying pesticide doses.

Several strengths of this study should be considered. The longitudinal burst design provided a large number of repeated cognitive observations from nearly 500 participants across multiple periods of high and low pesticide application. Mobile cognitive assessment allowed performance to be measured repeatedly in participants’ daily environments rather than relying exclusively on infrequent clinic-based neuropsychological assessments. This approach provides an opportunity to characterize short-term variability in cognition and to examine how performance changes in relation to temporally varying environmental and contextual factors. The inclusion of multiple cognitive tasks also allowed us to evaluate whether associations were consistent across cognitive domains.

These findings should also be interpreted in light of several limitations. First, pesticide spray season is an ecological proxy for periods of greater pesticide use and does not quantify individual pesticide exposure. Substantial variability in exposure may exist within a given spray season based on occupation, residential location, behaviors, and specific pesticide applications. Such exposure misclassification could attenuate associations with cognitive performance.

Second, floricultural employment similarly represents a proxy for a complex set of occupational exposures and conditions and cannot isolate the effects of pesticides from other workplace factors. Third, although the large number of repeated observations improves precision, repeated administration introduces the potential for practice effects, fatigue, and changes in engagement over time, which may remain despite our efforts to control for these through enhanced participant engagement and statistical modelling. The differing longitudinal trajectories across cognitive tasks emphasize the importance of these effects. An additional unresolved question is whether the relatively small, short-term fluctuations in cognitive performance observed across repeated assessments have implications for longer-term cognitive health. Although performance remained within the normative range in this cognitively normal young adult sample, repeated short-term changes in cognition could potentially signal vulnerability to later decline. Longitudinal follow-up will be needed to determine whether these fluctuations are transient and clinically inconsequential or are associated with longer-term cognitive trajectories.

In conclusion, this study reveals a complex dynamic between agricultural environments and neurobehavioral outcomes: while specific seasonal pesticide exposures may transiently stimulate certain cognitive domains, direct occupational exposure presents consistent cognitive risks. Peak pesticide spray seasons were associated with enhanced performance on specific processing speed tasks rather than broader cognitive changes. In contrast, direct floricultural employment was associated with significantly poorer performance across multiple cognitive measures, particularly in working memory and processing speed domains. These findings highlight the importance of evaluating both seasonal and occupational factors and demonstrate the complexity of short-term environmental influences on cognition. Future studies integrating intensive longitudinal cognitive assessment with individual-level pesticide biomarkers and detailed occupational exposure measures may help clarify the mechanisms underlying the cognitive differences observed among agricultural workers.

## Data Availability

Primary data analysis addressing the study aims are ongoing, and data is not just publicly available.

